# Advancing precision medicine in the Cardiac Intensive Care Unit using universal whole-genome sequencing

**DOI:** 10.64898/2026.05.11.26352916

**Authors:** Genevieve Kierulf, Melanie Emmerson, Patti Krautscheid, Steven B Bleyl, Martin Tristani-Firouzi, Briana L. Sawyer

## Abstract

Congenital heart defects (CHD) are a common congenital anomaly and a leading cause of neonatal mortality. Even in ostensibly isolated cases, genetic testing can reveal monogenic causes of isolated CHD or identify syndromic conditions before additional features become clinically apparent. A timely and accurate genetic diagnosis can inform medical management and surveillance, reduce the need for unnecessary investigations, and offer families valuable information about prognosis, recurrence risk, and anticipatory guidance. In September of 2023, Primary Children’s Hospital introduced a universal genetic testing protocol that implemented whole genome sequencing for all neonates admitted to the cardiac intensive care unit (CICU) undergoing cardiac surgery before 30 days of life, with the goal of increasing the number of patients who receive a timely genetic diagnosis and improving clinical care. This is a retrospective chart review of patients who underwent whole genome sequencing (WGS) under the new universal genetic testing protocol at Primary Children’s Hospital from its initiation in September 2023 to February 2026. Over the study period, 217 neonates with CHD participated in the universal WGS protocol. Of these patients, 23 (10.6%) received a genetic diagnosis that was causative of their CHD, of which 11 patients (48%) had no major extracardiac features at the time testing was ordered. Twenty patients were diagnosed with a syndromic condition, and three patients were diagnosed with a non-syndromic condition. All of these patients received additional referrals to specialists following their new diagnosis, and six families used results to inform decisions regarding continuation of care. An additional 19 patients (8.8%) received WGS results that were clinically relevant but non-diagnostic for their CHD, including partial diagnoses, secondary findings, and carrier status. In total, 19.4% of patients (n=42) had clinically relevant variants identified on their WGS.

## Introduction

Congenital heart defects (CHD) are a common congenital anomaly and a leading cause of neonatal mortality. CHDs are estimated to affect around 1% of births worldwide and approximately 20-40% of cases can be attributed to an identifiable genetic cause (Ahrens-Nicklas *et al*., 2016; Durbin *et al*., 2023; Helm and Ware, 2024). Twenty percent of CHDs are accompanied by extra cardiac anomalies, while 80% remain isolated (Wilde *et al*., 2022a).

Early genetic testing and evaluation are increasingly recognized as essential components of clinical care for children with CHD. A timely and accurate genetic diagnosis can inform medical management and surveillance, reduce the need for unnecessary investigations, and offer families valuable information about prognosis, recurrence risk, and anticipatory guidance. While genetic testing in CHD has traditionally been reserved for individuals with extracardiac anomalies or specific lesions that have a higher association with genetic conditions, a recent consensus statement recommended chromosomal microarray (CMA), at minimum, for all neonates with complex heart defects requiring intervention (Wilde *et al*., 2022b). Similarly, guidelines published by the National Society of Genetic Counselors (NSGC) recommend CMA as a first tier test for patients with both syndromic and non-syndromic CHDs (Ison *et al*., 2022). In addition, studies have shown that genetic testing based on the presence of extracardiac anomalies alone missed 13.4% of patients with an identifiable genetic cause suggesting utility in testing all patients with CHD (Helm and Ware, 2024).

In recent years, the cost of whole genome sequencing (WGS) has decreased, making it accessible as a first-tier test for patients presenting with CHDs. WGS has advantages over other genetic testing methodologies because it can simultaneously detect both copy number and single-nucleotide variants in a single test. In the CHD population, this type of broad testing modality is important as genetic CHD can have multiple underlying causes including aneuploidy, CNV, and single-nucleotide variants in growing number of genes. Utilizing WGS increases diagnostic yield over the currently recommended CMA given the genetic heterogeneity underlying CHD (Wilde *et al*., 2022b; Durbin *et al*., 2024; Shreeve *et al*., 2024).

To date, studies evaluating the diagnostic yield of WGS in CHD populations have largely focused on selected cohorts referred for testing based on clinical suspicion of an underlying genetic condition. Within these selected populations, WGS has identified actionable findings in approximately 27-46% of cases (Sweeney et al., 2021; Hays et al., 2023; Keefe et al., 2026). Furthermore, results from WGS have been shown to inform surgical planning in infants with CHD and significantly reduce hospitalization costs (Sweeney *et al*., 2021). However, the diagnostic yield and clinical utility of WGS in a broad, unselected CHD population remains relatively unknown. Notably, these prior studies primarily included patients referred for inpatient genetic counseling rather than evaluating a universal testing strategy.

Even in cases that appear to be isolated, genetic testing can reveal monogenic causes or identify syndromic conditions before additional features become clinically apparent. While diagnostic yield is generally lower in isolated or seemingly non-syndromic cases, some institutions adopt a broader or more universal strategy to enable earlier identification of genetic conditions that may inform prognosis, medical management and surgical decision making, potentially before extracardiac features become clinically apparent. Supporting this approach, a recent study found that genetic testing with a broad CHD panel and exome-wide CNV analysis identified a diagnosis in 14% of isolated CHD patients (Rajan *et al*., 2026).

Despite these benefits, the approach to implementing genetic counseling and/or testing for appropriate patients often varies significantly between institutions (Callahan *et al*., 2022, 2023). Studies highlight a persistent gap between the recognized utility of genetic testing and barriers to its implementation in the acute care setting, such as differences in institutional protocols, resource availability, and under-recognition of possible genetic conditions (Char *et al*., 2018; Callahan *et al*., 2022; Wojcik, Del Rosario and Agrawal, 2022).

In September of 2023, Primary Children’s Hospital introduced a new protocol that implemented universal clinical WGS for all neonates admitted to the cardiac intensive care unit undergoing cardiac surgery. The goal of this protocol was to increase the number of patients who receive an early genetic diagnosis to support timely clinical care. Identifying an underlying genetic cause can guide clinical management decisions, inform prognosis, identify other at-risk family members, and provide estimates of recurrence risk for the family. In addition, non-diagnostic WGS results can provide reassurance to families and providers against the broad differential of genetic syndromes associated with CHD. The goal of this study is to evaluate the implementation of this universal WGS protocol in patients with an ostensibly isolated heart defect in the neonatal period.

The specific aims of the study are as follows: 1. To determine the percentage of patients with CHDs who received a diagnosis under our institution’s universal WGS protocol; 2. To compare the diagnostic yield among patients with isolated CHD to patients with extracardiac anomalies at the time of testing; and 3. To evaluate how WGS has impacted clinical management and family counseling for patients with CHDs.

## 1 Methods

This is a retrospective chart review of neonates who underwent a universal WGS protocol at Primary Children’s Hospital from September 2023 to February 2026. This study was approved by the University of Utah’s Institutional Review Board and was determined to be exempt from human subjects review. Patients admitted to the CICU under 30 days of life with a CHD that may require surgical repair in infancy were offered clinical WGS following consultation and consent with a genetic counselor (Figure 1). Patients were identified for genetic counseling consultation based on an automatic alert through the electronic medical record (EMR). In cases where CHD was diagnosed prenatally, parents had the option to receive genetic counseling and consent to postnatal testing in the fetal period. Patients with known or highly suspected aneuploidy condition (e.g. trisomy 13, 18, 21) based on prenatal screening, prenatal diagnostic testing, or ultrasound findings were excluded from WGS and not included in this study. CHD patients admitted to the neonatal intensive care unit for prematurity or other pertinent health concerns are not included in this cohort if they transferred to the CICU after 30 days of life or already had genetic testing prior to transfer.

**Figure 1.**
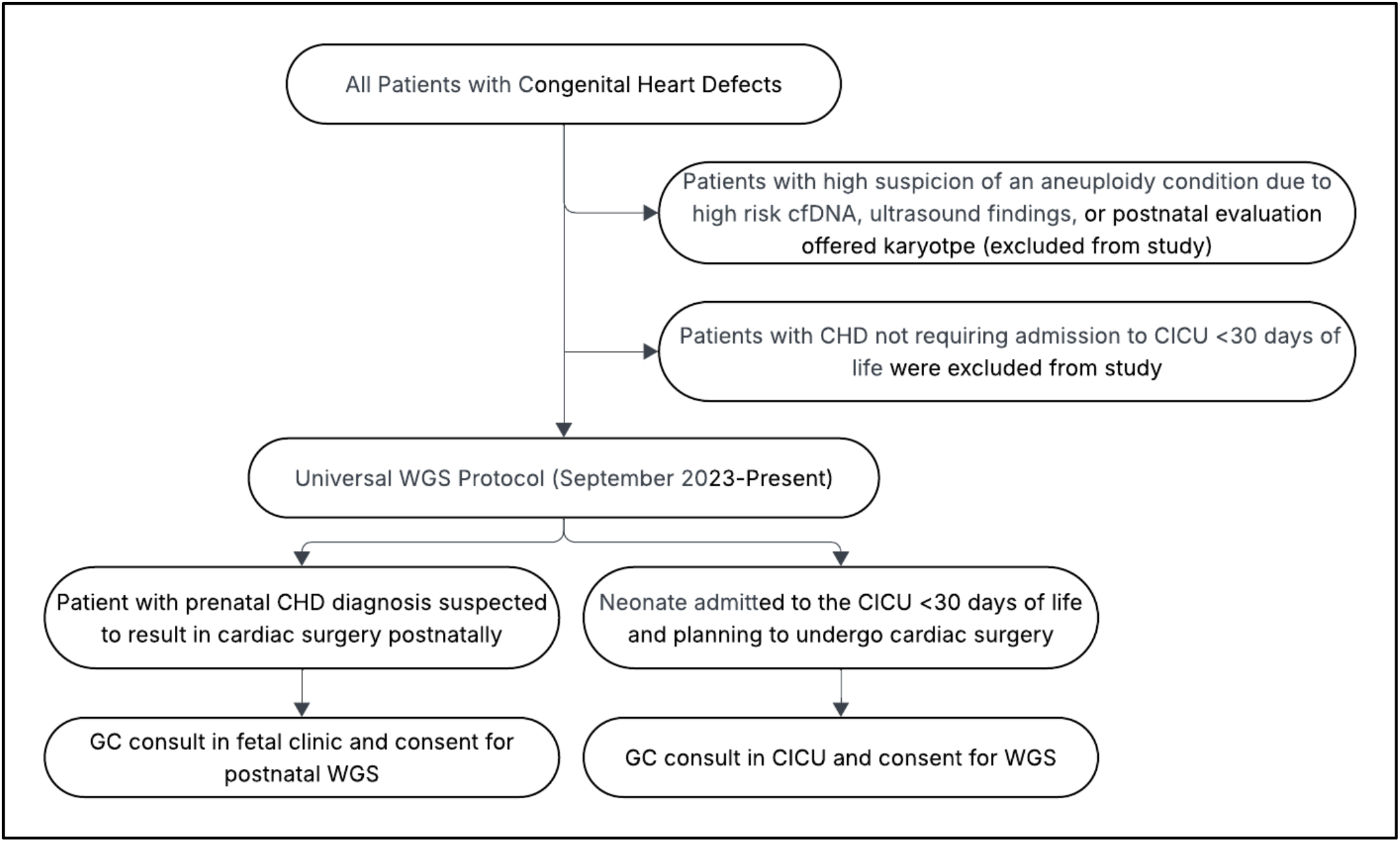
Genetic Testing Protocol. This flow chart outlines the universal WGS protocol implemented in September 2023 compared to our institution’s previous lesion-specific protocol. Abbreviations: CHD, congenital heart defect; WGS, whole genome sequencing; CICU, cardiac intensive care unit.

Under the universal WGS protocol, patients were offered non-rapid, proband-only WGS as the default testing option. Trio whole genome sequencing was offered when a family history of CHD was present to aid in result interpretation. Trio rapid and ultra-rapid testing was offered for patients who were either acutely ill, had extracardiac anomalies, or where providers expressed preference to have results for surgical decision-making. While testing typically followed the strategy outlined in Figure 1, testing was recommended individually considering clinical context and provider recommendations.

Clinical data were obtained from the electronic medical record and collected in REDCap (Research Electronic Data Capture), a secure, web-based software designed to support clinical research.

Additional data abstracted and analyzed from medical records included the following: demographics, type of CHD, extracardiac anomalies, genetic diagnosis, type of variant detected, variant interpretation, phenotypic terms included on the genetic test report, secondary/incidental findings, family/prenatal history, referrals to specialists, procedures and surgical history, and follow-up care.

Sequencing and variant interpretation were performed by commercial CLIA-certified genetic testing laboratories (Broad Institute, Rady Children’s Hospital, GeneDx, and ARUP). Variants identified on WGS were considered diagnostic if they were classified as pathogenic or likely pathogenic according to American College of Medical Genetics (ACMG) guidelines and deemed causative of the patient’s congenital heart disease (CHD). Pathogenic variants explaining features of a patient’s phenotype unrelated to CHD were considered a partial diagnosis. Variants classified as uncertain significance by the testing laboratory underwent additional review by the clinical genetics team to assess concordance with the patient’s evolving phenotype. In select cases, variants of uncertain significance with strong clinical correlation that directly informed clinical management were considered diagnostic. All other variants of uncertain significance, including those with suggestive but insufficient evidence, were classified as non-diagnostic for the purposes of this analysis.

## 2 Results

Over the study period, 217 neonates with CHD participated in the universal WGS protocol. The majority of the study population was male (59.4%, n=129). Thirteen percent of patients (n=28) passed away during the follow-up period. The median age at the time that genetic testing was initiated was 2.8 days of life. Genetic testing included proband-only non-rapid WGS (70.5%, n=153), rapid duo/trio WGS (17.5%, n=38), rapid proband-only WGS (7.4%, n=16), and duo/trio non-rapid WGS (4.6%, n=10) (Table 1). In total, 19.4% of patients (n=42) had clinically relevant variants identified on their WGS (Figure 2).

**Table 1.** Demographics

| Variable (n=217) | Value (%) |
| --- | --- |
| Gender |  |
| Male | 129 (59.4%) |
| Female | 87 (40.1%) |
| Median age at time of testing | 2 days of life |
| Deceased | 28 (12.9%) |
| Genetic Testing |  |
| Proband Only WGS | 153 (70.5%) |
| Duo/Trio WGS | 10 (4.6%) |
| Proband Only WGS (Rapid) | 16 (7.4%) |
| Duo/Trio WGS (Rapid) | 38 (17.5%) |

**Figure 2.**
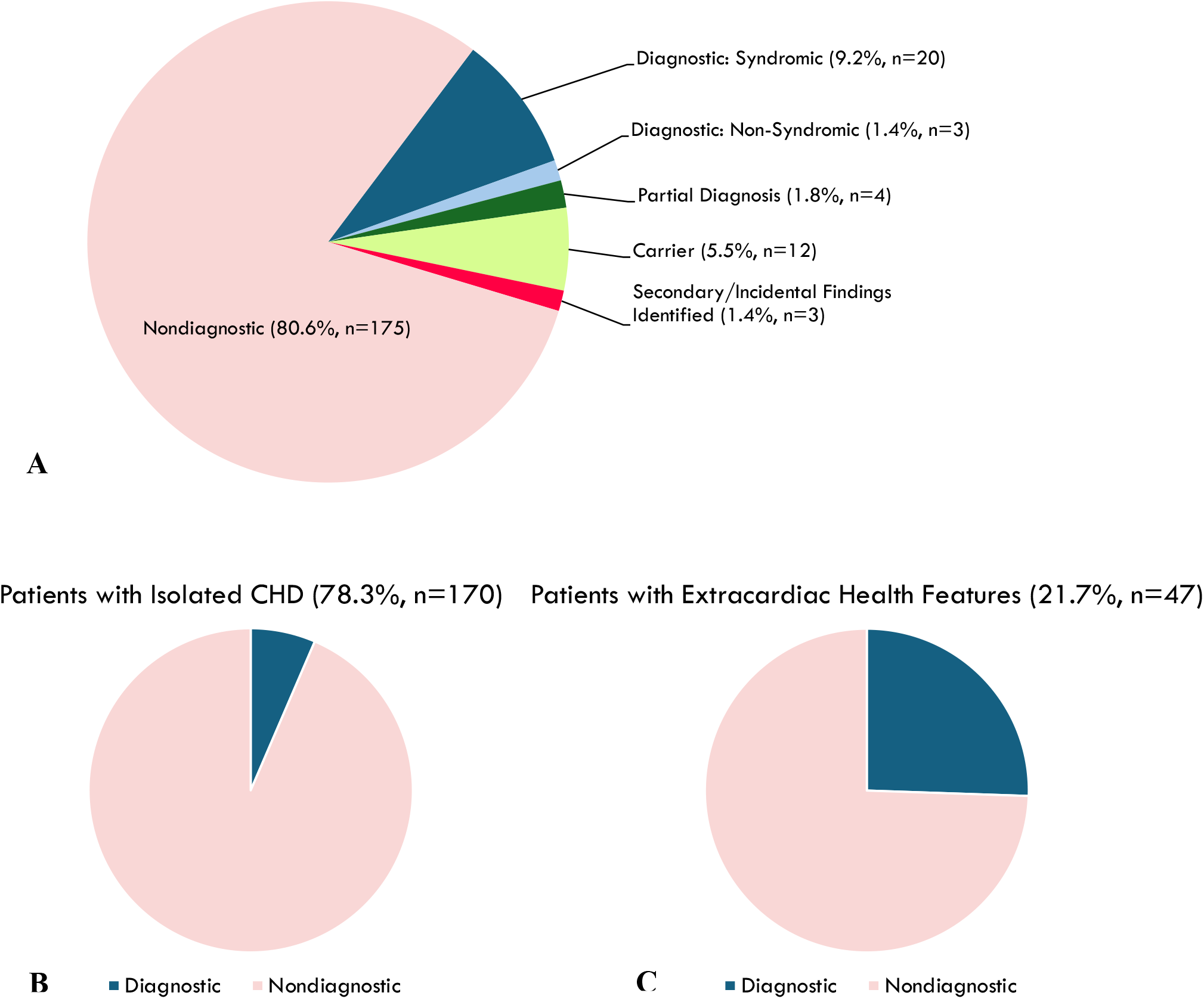
Overall Yield of WGS Protocol. Figure 2. Panel A illustrates the overall yield of the universal WGS protocol that was implemented in September of 2023. Panels B and C compare the diagnostic yield of WGS for patients with an isolated CHD versus those with extracardiac anomalies at the time of testing.

### Overall Yield of Universal WGS

Overall, 23 patients (10.6%) received a genetic diagnosis that explained the etiology of their CHD. Twenty patients were diagnosed with a syndromic condition, and three patients were diagnosed with a non-syndromic cause of their CHD. Syndromic diagnoses included 22q11.2 Deletion Syndrome (n=4), William’s Syndrome (n=2), Capillary Alveolar Dysplasia (n=2), Alagille Syndrome (n=2), Zellweger Syndrome, CHARGE Syndrome, Kabuki Syndrome, Holt Oram Syndrome, 1q21.2 Duplication Syndrome, 15q13.3 Deletion Syndrome, Cat Eye Syndrome, Noonan Syndrome, Partial Trisomy 13/Wolff-Hirschhorn Syndrome, and an unbalanced translocation involving 18q and 4q. Patients with non-syndromic diagnoses had pathogenic variants identified in *FLT4, NOTCH1*, and *ZFPM2* (Table 2).

**Table 2.**
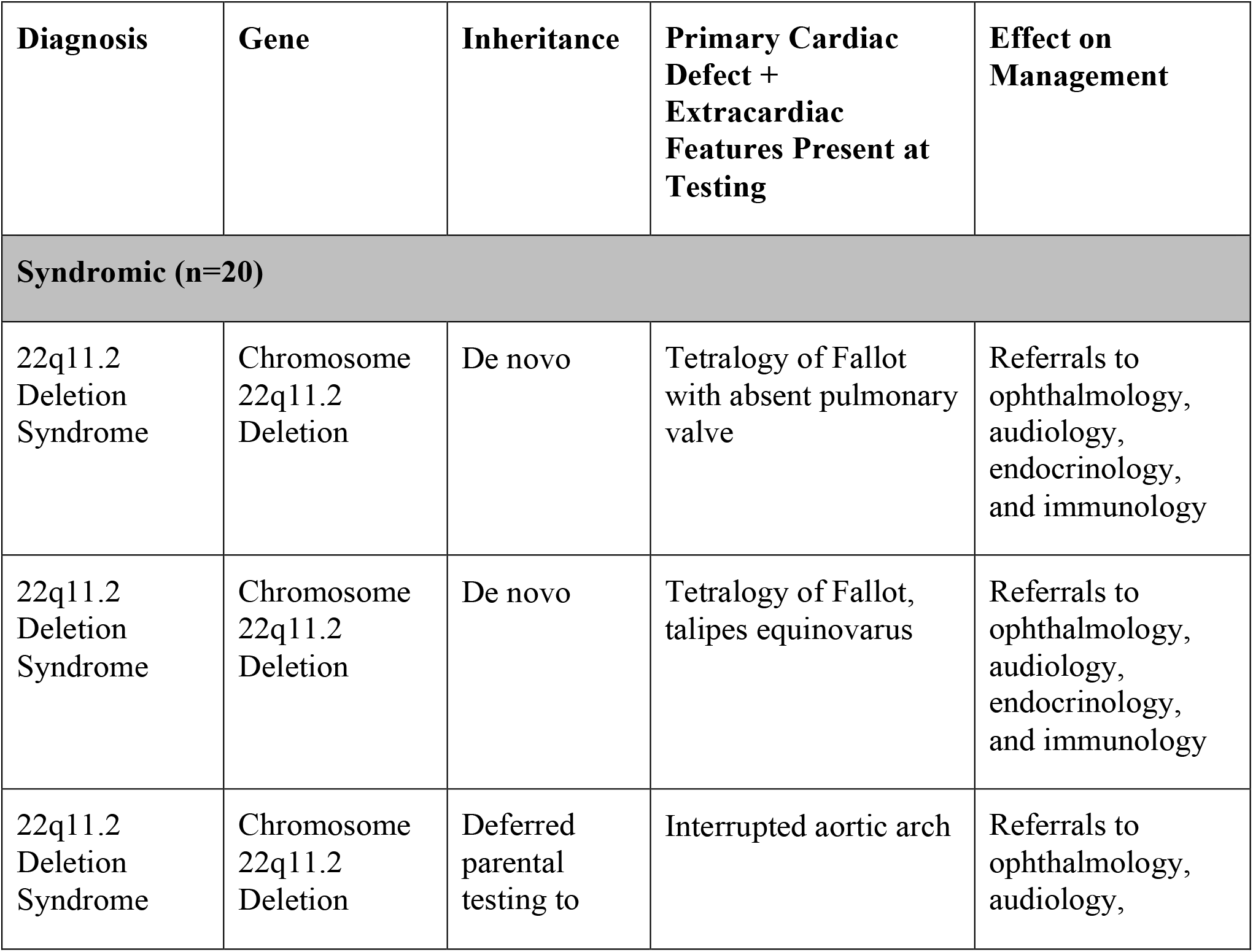
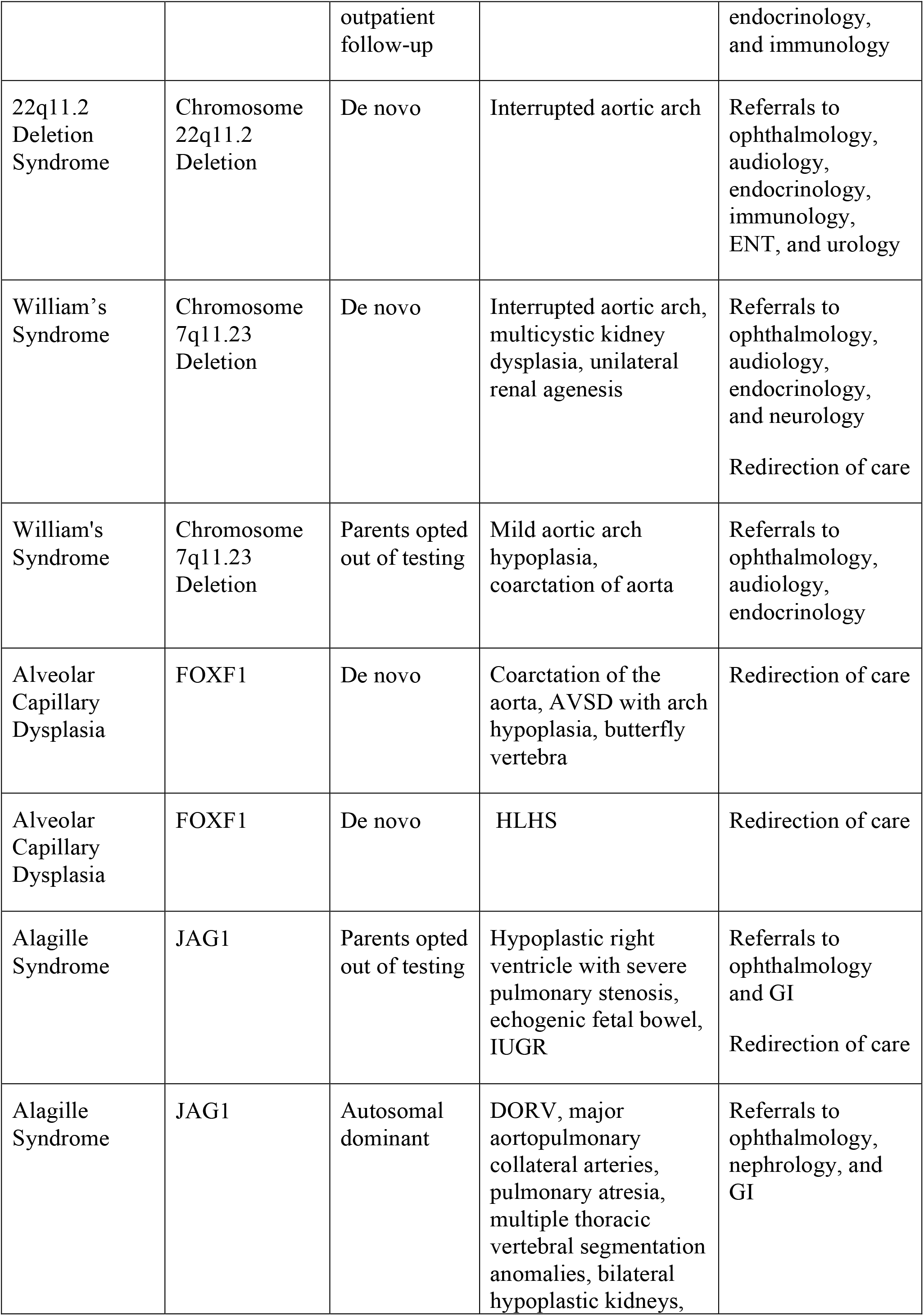
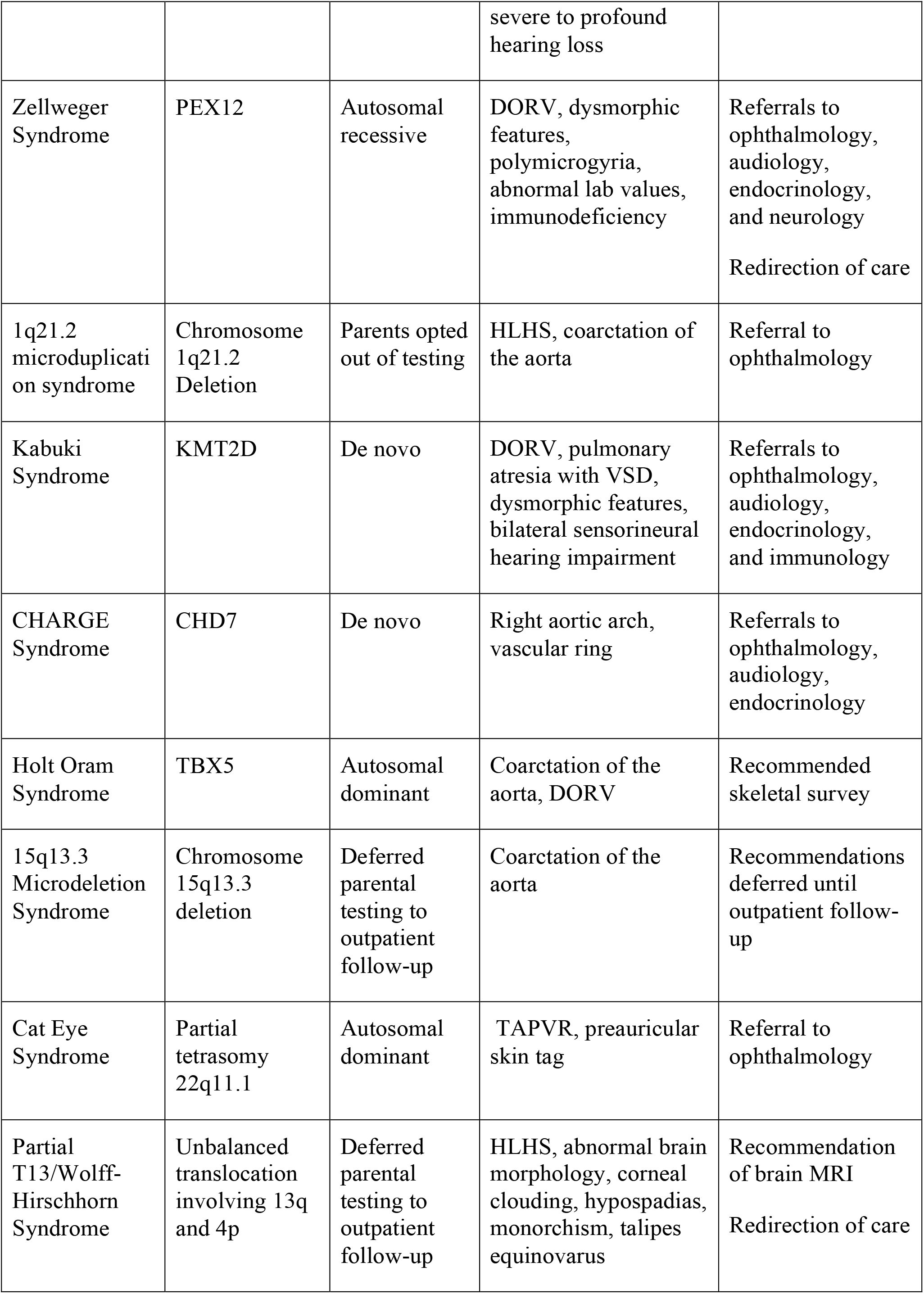
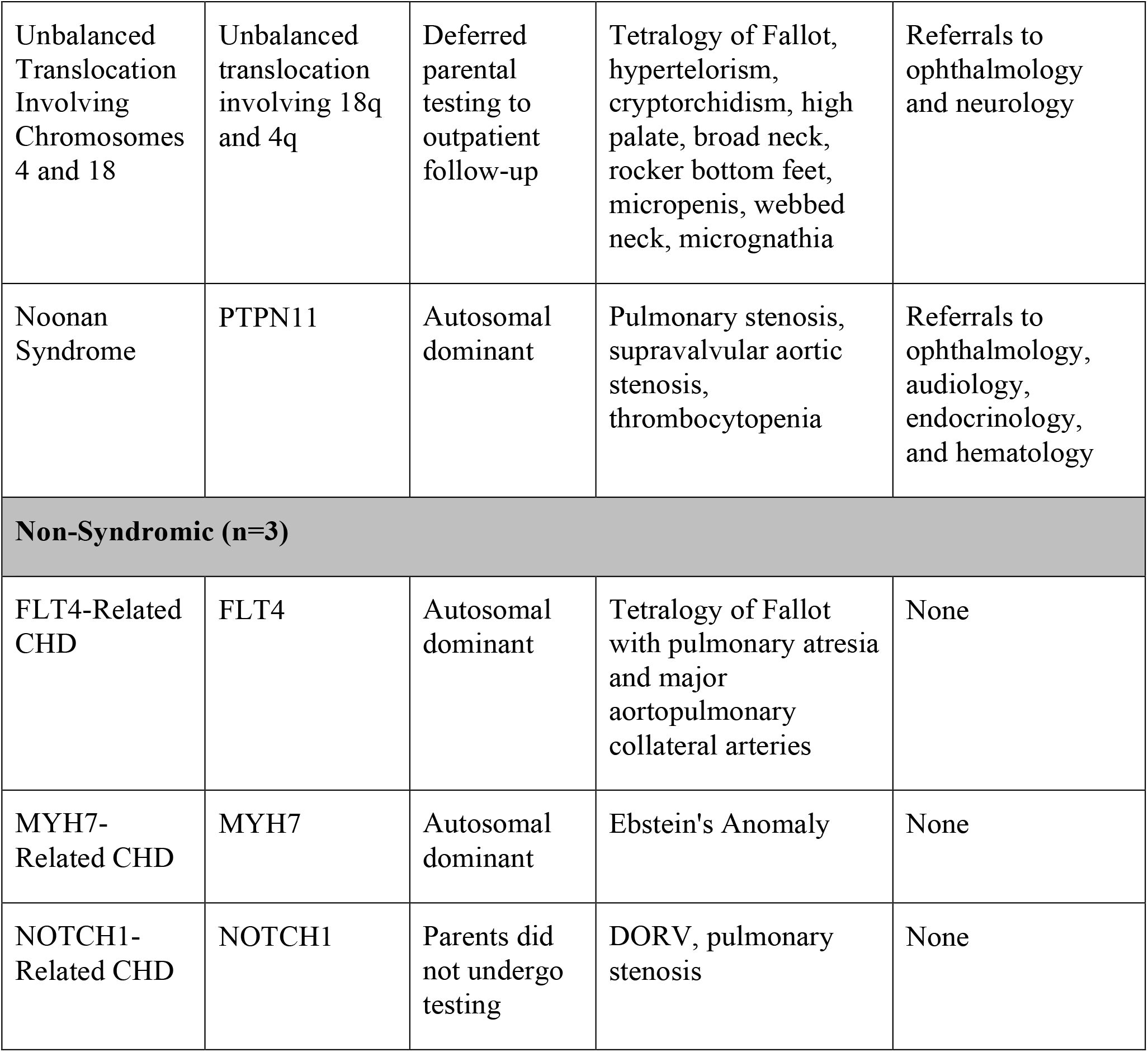
Genetic Diagnosis and Effect on Management

In addition, 19 patients (8.8%) received WGS results that were clinically relevant but non-diagnostic for their CHD. Four patients (1.8%) received a partial diagnosis that explained aspects of their phenotype unrelated to their heart defect, including Sickle Beta Plus Thalassemia (*HBB*), Prothrombin Deficiency (*F2*), COL4A1-related small vessel vasculopathy (*COL4A1*), and an Intellectual Disability-Severe Speech Delay-Mild Dysmorphism Syndrome (*FOXP1*). Three patients (1.4%) had secondary findings identified at the time of testing, including Familial Hypercholesterolemia (*LDLR*), Nonsyndromic Hypertrophic Cardiomyopathy (*MYH7*), and Butylcholinesterase Deficiency (*BCHE*). Carrier status was identified in twelve patients (5.5%). Overall, 19.4% of patients had clinically relevant variants identified on WGS (Figure 2).

In total, 39 patients (18%) had 47 variants of uncertain significance reported. These were reviewed by the study team to assess variant characteristic and phenotypic fit. One patient had a VUS with strong enough clinical correlation and suspicion that it was considered a diagnostic result and used in management. The remainder of the VUSs that did not have good phenotypic overlap or remain unresolved were considered non-diagnostic results.

Prior to our universal WGS protocol, we would reserve genetic testing for lesions associated with a higher pre-test likelihood of a genetic diagnosis. For example, we would offer a chromosomal microarray for conotruncal defects due to their known association with 22q11.2 deletion syndrome. We reviewed our patients’ diagnoses and determined that eight of twenty-three diagnoses (35%) identified by WGS would likely not have been identified through our prior lesion-based testing approaches.

### Yield of WGS Based on Phenotype at Time of Testing

At the time of testing, 78.3% of patients (n=170) had no known extracardiac anomalies. Of these patients, 6.5% (n=11) were diagnosed with a genetic condition. Eight of these diagnoses were syndromic and three were non-syndromic (Table 2).

Conversely, there were 47 patients (21.7%) who had extracardiac features at time of test initiation. Twelve of these patients (25.5%) received a genetic diagnosis, all of which were syndromic. One of the patients with 22q11.2 deletion syndrome had high-risk cell free DNA screening during pregnancy that alluded to the diagnosis prior to WGS. Two patients with 22q11.2 deletion syndrome had low risk cfDNA that did not include screening for microdeletion syndromes, and one did not undergo cfDNA screening.

Analysis of the diagnostic yield based on phenotype found that patients with extracardiac anomalies at the time of testing were more likely to receive a genetic diagnosis compared to those with an isolated CHD (25.5% vs 6.5%, p <0.001). This difference remains significant when including patients with clinically relevant variants that were not causative of their CHD (31.9% vs 15.9%, p=0.014).

There were 194 patients who did not receive a genetic diagnosis. Thirty-four of these patients (17.5%) had extracardiac anomalies at the time of testing. An additional 6 initially had isolated CHD at the time of testing but developed extracardiac anomalies during their admission or shortly following discharge.

### Impact on Clinical Management Decisions and Family Counseling

Patients with a genetic diagnosis received additional referrals to specialists based on the expected clinical manifestations of their condition (Table 2). Patients received referrals for supportive therapies (n=21) and developmental assessments (n=11), as well as targeted referrals to specialties including ophthalmology (n=15), audiology (n=10), endocrinology (n=10), immunology (n=5), and neurology (n= 3) based on anticipated manifestations of their diagnoses. Thirteen patients with a syndromic diagnosis had additional imaging ordered as a result of their diagnosis, including renal ultrasounds (n=12), additional radiography (n=8), and brain MRIs (n=3). Families and care teams incorporated a patient’s genetic diagnosis and expected prognosis when evaluating next steps in care. Six families (26%) chose to redirect to a comfort care pathway following these discussions and incorporation of the diagnosis. Genetic diagnoses that contributed to redirection of care were Alveolar Capillary Dysplasia (n=2), Zellweger Syndrome, Alagille Syndrome, William’s Syndrome, and Partial T13/Wolff Hirschhorn Syndrome.

Of families that received a genetic diagnosis for their child, 69.6% (n=16) underwent parental testing to determine inheritance of the variant. Eight of these were included as a part of their child’s trio or duo WGS analysis, and an additional eight pursued targeted testing following the return of proband-only WGS results. Following this parental testing, eight variants were found to be de novo and seven inherited, six of which were autosomal dominant and one autosomal recessive. Three families (13%) opted out of parental testing, and four (17.4%) expressed interest but deferred testing until their outpatient follow-up visit.

## 3 Discussion

Implementation of universal WGS identified a genetic diagnosis in 10.6% of patients with a CHD, almost half of whom (48%) had no known extracardiac features at the time of testing. Diagnoses in our patient population included established microdeletion and duplication syndromes, in addition to various single-gene disorders. Of those that were diagnosed, eight patients would not have been offered testing under our institution’s prior targeted genetic testing protocol. Previously, patients with isolated heart defects that were not highly associated with a specific genetic condition were unlikely to undergo genetic testing until they presented with extracardiac manifestations later in their care. Implementing universal WGS enabled these patients to receive a genetic diagnosis early in life, which allowed for informed decision-making and more targeted clinical recommendations.

In addition to identifying causative variants for CHD, WGS identified clinically relevant variants in an additional 8.8% of patients, including partial diagnoses, secondary findings, and carrier status. These findings impacted clinical management by explaining other aspects of a patient’s phenotype, guiding surveillance recommendations, and informing reproductive risks.

Patients with extracardiac anomalies at the time of testing were more likely to receive a genetic diagnosis compared to those with an isolated CHD (25.5% vs 6.5%, p <0.001). This aligns with what has been reported in the literature for patients with CHD using targeted genetic testing protocols (Helm and Ware, 2024). We found that 15% of undiagnosed patients who initially presented with an isolated CHD went on to develop extracardiac anomalies during the follow-up period. An added benefit of utilizing WGS in this population is the ability to reanalyze sequencing data using new phenotype terms when additional health concerns develop over their lifetime compared to other genetic testing technologies.

Diagnostic yields of genetic testing in CHD populations have ranged from 20-30%, however, these studies have historically included patients who were referred by the clinical team following suspicion of a genetic condition or those with a syndromic presentation, resulting in a cohort enriched for patients more likely to receive a diagnosis. (Sweeney *et al*., 2021; Keefe *et al*., 2026). In contrast, our study provided an unbiased and accurate yield of genetic testing in an unselected group of patients with CHD. As genetic testing becomes more accessible, offering WGS as a first-tier test for patients with CHD helps to eliminate clinician bias and promotes equity within the healthcare system.

For patients who received a syndromic diagnosis, clinicians were able to better anticipate healthcare needs and provide appropriate referrals early on, which has been shown to positively impact long term outcomes for CHD patients (Simmons and Brueckner, 2017). Identifying a genetic diagnosis has also been shown to decrease overall healthcare costs by reducing unnecessary clinical workup and interventions as well as time spent in the hospital (Farnaes *et al*., 2018). In cases where families ultimately decided to redirect to a palliative care pathway, the clinical team incorporated the genetic diagnosis, impact of the patient’s CHD, and other associated health concerns when discussing surgical options with families. Following these discussions, families were able to incorporate information about the expected prognosis and life outcomes associated with their child’s condition when making decisions regarding continuation of care in collaboration with their care team.

Identifying a genetic diagnosis can provide clarity in the decision-making process and help to bring closure to families whose children have passed away.

For those who received a non-syndromic diagnosis, genetic testing allowed for improved recurrence risk counseling for families and helped to alleviate concern regarding potential extracardiac manifestations. In these cases, families could be reassured that their child was unlikely to develop additional health concerns related to their genetic diagnosis and clinicians could feel comfortable discharging patients without completing unnecessary workup.

Recent work has also demonstrated the prognostic value of genetic testing for outcomes prediction for specific types of CHD, mostly focusing on the impact of copy number variants (Carey *et al*., 2013; Kim *et al*., 2016; Boskovski *et al*., 2020; Geddes, Przybylowski and Ware, 2020; Landis *et al*., 2022). Beyond copy number variants, damaging genotypes in specific gene pathways were associated with an elevated risk of adverse post-operative outcomes, including mortality, cardiac arrest and prolonged mechanical ventilation (Watkins *et al*., 2025a). The impact of damaging genotypes was further amplified in the context of specific CHD phenotypes, surgical complexity and extra-cardiac anomalies. The absence of a damaging genotype in these gene pathways was also informative, reducing the risk of adverse postoperative outcomes (Watkins *et al*., 2025b). Taken together, these studies demonstrate that genome sequencing may enable outcomes prediction and risk stratification following congenital cardiac surgery.

There are several limitations to this study that likely contribute to the differences seen in diagnostic yield when compared to other studies. First, our study population does not include patients who were diagnosed prenatally or received targeted testing at birth following high risk cfDNA screening, which effectively excludes any patients with an aneuploidy condition. Our study population also does not include patients with a heart defect that did not require surgical intervention or admission to the cardiac ICU, as these patients were typically not offered genetic counseling consultation unless they presented with extracardiac anomalies. Additionally, several patients who were recently diagnosed had limited clinical follow-up prior to February 2026, which may have resulted in an incomplete analysis of the impact on clinical management and decision-making.

As a result of this study, additional research could seek to contextualize the proportion of patients included in our study population within all those diagnosed with CHD in order to develop a more well-rounded understanding of the genetic etiology of CHD. Future research could also investigate the yield on reanalysis of WGS data as genetic knowledge and patients’ phenotypes continue to evolve. Lastly, studies could quantitatively examine the impact on resource utilization and cost reduction within the healthcare system resulting from targeted management strategies.

## 4 Conclusions

In summary, our institution’s universal genetic testing protocol demonstrated the utility of WGS in patients with isolated congenital heart defects. In total, 19.4% of patients received clinically relevant results on their WGS, including many who would not have previously been offered genetic testing.

Receiving a genetic diagnosis guided clinical management by informing decisions regarding continuation of care and enabling these patients to receive appropriate referrals to specialists. Additionally, diagnostic WGS results allowed for more accurate counseling on recurrence risks for future pregnancies.

## Data Availability

The original contributions presented in the study are included in the article/supplementary material, further inquiries can be directed to the corresponding author/s.

## 6 Author Contributions

G.K: Conceptualization, methodology, investigation, data curation, formal analysis, visualization, writing—original draft, writing—review and editing

M.E.: Conceptualization, methodology, investigation, supervision, resources, writing—review and editing

B.S.: Conceptualization, methodology, investigation, supervision, resources, writing—review and editing

P.K.: Conceptualization, supervision, writing—review and editing

S.B.: Writing—review and editing

M.T.F.: Writing—review and editing

## References

Ahrens-Nicklas, R.C. et al. (2016) “Utility of genetic evaluation in infants with congenital heart defects admitted to the cardiac intensive care unit,” American Journal of Medical Genetics. Part A, 170(12), pp. 3090–3097. Available at: 10.1002/ajmg.a.37891.

Boskovski, M.T. et al. (2020) “De Novo Damaging Variants, Clinical Phenotypes, and Post-Operative Outcomes in Congenital Heart Disease,” Circulation. Genomic and Precision Medicine, 13(4), p. e002836. Available at: 10.1161/CIRCGEN.119.002836.

Callahan, K.P. et al. (2022) “How neonatologists use genetic testing: findings from a national survey,” Journal of Perinatology: Official Journal of the California Perinatal Association, 42(2), pp. 260–261. Available at: 10.1038/s41372-021-01283-4.

Callahan, K.P. et al. (2023) “Hospital-level variation in genetic testing in children’s hospitals’ neonatal intensive care units from 2016 to 2021,” Genetics in Medicine: Official Journal of the American College of Medical Genetics, 25(3), p. 100357. Available at: 10.1016/j.gim.2022.12.004.

Carey, A.S. et al. (2013) “Effect of copy number variants on outcomes for infants with single ventricle heart defects,” Circulation. Cardiovascular Genetics, 6(5), pp. 444–451. Available at: 10.1161/CIRCGENETICS.113.000189.

Char, D.S. et al. (2018) “Anticipating uncertainty and irrevocable decisions: provider perspectives on implementing whole-genome sequencing in critically ill children with heart disease,” Genetics in Medicine: Official Journal of the American College of Medical Genetics, 20(11), pp. 1455–1461. Available at: 10.1038/gim.2018.25.

Durbin, M.D. et al. (2023) “A multicenter cross-sectional study in infants with congenital heart defects demonstrates high diagnostic yield of genetic testing but variable evaluation practices,” Genetics in Medicine Open, 1(1), p. 100814. Available at: 10.1016/j.gimo.2023.100814.

Durbin, M.D. et al. (2024) “Rapid Genome Sequencing Shows Diagnostic Utility In Infants With Congenital Heart Defects,” Research Square, p. rs.3.rs-3976548. Available at: 10.21203/rs.3.rs-3976548/v1.

Farnaes, L. et al. (2018) “Rapid whole-genome sequencing decreases infant morbidity and cost of hospitalization,” NPJ genomic medicine, 3, p. 10. Available at: 10.1038/s41525-018-0049-4.

Geddes, G.C., Przybylowski, L.F. and Ware, S.M. (2020) “Variants of significance: medical genetics and surgical outcomes in congenital heart disease,” Current Opinion in Pediatrics, 32(6), pp. 730–738. Available at: 10.1097/MOP.0000000000000949.

Helm, B.M. and Ware, S.M. (2024) “Clinical Decision Analysis of Genetic Evaluation and Testing in 1013 Intensive Care Unit Infants with Congenital Heart Defects Supports Universal Genetic Testing,” Genes, 15(4), p. 505. Available at: 10.3390/genes15040505.

Ison, H.E. et al. (2022) “Genetic counseling for congenital heart disease - Practice resource of the National Society of Genetic Counselors,” Journal of Genetic Counseling, 31(1), pp. 9–33. Available at: 10.1002/jgc4.1498.

Keefe, A.C. et al. (2026) “Implementation of First-Line Rapid Genome Sequencing for Children in Pediatric and Cardiac Intensive Care Units,” American Journal of Medical Genetics. Part A [Preprint]. Available at: 10.1002/ajmg.a.70088.

Kim, D.S. et al. (2016) “Burden of potentially pathologic copy number variants is higher in children with isolated congenital heart disease and significantly impairs covariate-adjusted transplant-free survival,” The Journal of Thoracic and Cardiovascular Surgery, 151(4), pp. 1147–1151.e4. Available at: 10.1016/j.jtcvs.2015.09.136.

Landis, B.J. et al. (2022) “Learning to Crawl: Determining the Role of Genetic Abnormalities on Postoperative Outcomes in Congenital Heart Disease,” Journal of the American Heart Association, 11(19), p. e026369. Available at: 10.1161/JAHA.122.026369.

Rajan, R. et al. (2026) “Uptake of genetic testing for infants with congenital heart disease: Impact of prenatal versus postnatal cardiovascular genetic counseling,” Journal of Genetic Counseling, 35(2), p. e70194. Available at: 10.1002/jgc4.70194.

Shreeve, N. et al. (2024) “Incremental yield of whole-genome sequencing over chromosomal microarray analysis and exome sequencing for congenital anomalies in prenatal period and infancy: systematic review and meta-analysis,” Ultrasound in Obstetrics & Gynecology: The Official Journal of the International Society of Ultrasound in Obstetrics and Gynecology, 63(1), pp. 15–23. Available at: 10.1002/uog.27491.

Simmons, M.A. and Brueckner, M. (2017) “The genetics of congenital heart disease… understanding and improving long-term outcomes in congenital heart disease: a review for the general cardiologist and primary care physician,” Current Opinion in Pediatrics, 29(5), pp. 520–528. Available at: 10.1097/MOP.0000000000000538.

Sweeney, N.M. et al. (2021) “Rapid whole genome sequencing impacts care and resource utilization in infants with congenital heart disease,” NPJ genomic medicine, 6(1), p. 29. Available at: 10.1038/s41525-021-00192-x.

Watkins, W.S. et al. (2025a) “Genome sequencing is critical for forecasting outcomes following congenital cardiac surgery,” Nature Communications, 16(1), p. 6365. Available at: 10.1038/s41467-025-61625-0.

Watkins, W.S. et al. (2025b) “Genome sequencing is critical for forecasting outcomes following congenital cardiac surgery,” Nature Communications, 16(1), p. 6365. Available at: 10.1038/s41467-025-61625-0.

Wilde, A.A.M. et al. (2022a) “European Heart Rhythm Association (EHRA)/Heart Rhythm Society (HRS)/Asia Pacific Heart Rhythm Society (APHRS)/Latin American Heart Rhythm Society (LAHRS) Expert Consensus Statement on the state of genetic testing for cardiac diseases,” Europace: European Pacing, Arrhythmias, and Cardiac Electrophysiology: Journal of the Working Groups on Cardiac Pacing, Arrhythmias, and Cardiac Cellular Electrophysiology of the European Society of Cardiology, 24(8), pp. 1307–1367. Available at: 10.1093/europace/euac030.

Wilde, A.A.M. et al. (2022b) “European Heart Rhythm Association (EHRA)/Heart Rhythm Society (HRS)/Asia Pacific Heart Rhythm Society (APHRS)/Latin American Heart Rhythm Society (LAHRS) Expert Consensus Statement on the state of genetic testing for cardiac diseases,” Europace: European Pacing, Arrhythmias, and Cardiac Electrophysiology: Journal of the Working Groups on Cardiac Pacing, Arrhythmias, and Cardiac Cellular Electrophysiology of the European Society of Cardiology, 24(8), pp. 1307–1367. Available at: 10.1093/europace/euac030.

Wojcik, M.H., Del Rosario, M.C. and Agrawal, P.B. (2022) “Perspectives of United States neonatologists on genetic testing practices,” Genetics in Medicine: Official Journal of the American College of Medical Genetics, 24(6), pp. 1372–1377. Available at: 10.1016/j.gim.2022.02.009.

